# Test negative designs with uncertainty, sensitivity, and specificity

**DOI:** 10.1101/2021.06.24.21259495

**Authors:** Erik K. Johnson, Rebecca Kahn, Yonatan H. Grad, Marc Lipsitch, Daniel B. Larremore

**Affiliations:** Department of Applied Mathematics, University of Colorado Boulder, Boulder, CO, USA; Center for Communicable Disease Dynamics, Department of Epidemiology, Harvard T.H. Chan School of Public Health, Boston, MA, USA; Department of Immunology and Infectious Diseases, Harvard T.H. Chan School of Public Health, Boston, MA, USA; Department of Epidemiology, Harvard T.H. Chan School of Public Health, Boston, MA, USA; Department of Computer Science, University of Colorado Boulder; BioFrontiers Institute, University of Colorado Boulder

## Abstract

Test-negative designs (TNDs) can be used to estimate vaccine effectiveness by comparing the relative rates of the target disease and symptomatically similar diseases among vaccinated and unvaccinated populations. However, the diagnostic tests used to identify the target disease typically suffer from imperfect sensitivity and specificity, leading to biased vaccine effectiveness estimates. Here we present a solution to this problem via a Bayesian statistical model which can either incorporate point estimates of test sensitivity and specificity, or can jointly infer them directly from laboratory validation data. This approach enables uncertainties in the performance characteristics of the diagnostic test to be correctly propagated to estimates, avoiding both bias and false precision in vaccine effectiveness. By further incorporating individual covariates of study participants, and by allowing data streams from multiple diagnostic test types to be rigorously combined, our approach provides a flexible model for the analysis of TNDs with explicitly stated assumptions.

## Introduction

Test-negative designs (TNDs) have become a common design to study vaccine effectiveness [1, 2]. In TNDs, the study population consists of individuals who present to a medical clinic with symptoms resembling those of the disease of interest. We call the disease of interest the “target disease” or “target infection” depending on the endpoint of interest and we call symptomatically similar diseases “non-target diseases”. In TNDs, vaccine effectiveness is estimated by comparing the number of vaccinated patients who test positive and negative for the target disease to the number of unvaccinated patients who test positive and negative for the target disease.

Unfortunately, an imperfect diagnostic test may result in false positives (if specificity is imperfect) and false negatives (if sensitivity is imperfect). In turn, these false positives or false negatives, alternatively called outcome misclassifications, will lead to biased estimates of vaccine effectiveness unless statistical corrections are made [3, 4, 5, 6, 7]. Thus, appropriate statistical consideration of imperfect diagnostic tests, as well as potential confounders, is important for vaccine effective estimation and public health decision making.

Aspects of this problem have been studied by others. For instance, Endo et al. [4] proposed a frequentist bias-correction method for TNDs that accounts for imperfect sensitivity and specificity along with potential confounders (e.g., age and sex). Their method assumes that the true sensitivity and specificity of the diagnostic test are known, yet in practice, these values are not known exactly because they are, themselves, estimated from data. In the context of SARS-CoV-2 seroprevalence studies, failure to incorporate such uncertainty in sensitivity and specificity has led to incorrect conclusions [8, 9]. More broadly, small discrepancies between the assumed and actual sensitivity and specificity values can produce large biases [10, 11].

Jackson et al. [6] compared a Bayesian approach and a frequentist approach for estimating vaccine effectiveness in TNDs, but without consideration of imperfect test sensitivity and specificity. Imperfect tests have been analyzed in Bayesian frameworks, however, in the context of case-control studies in pertussis [12] and more general prevalence estimation [9, 13]. Thus, at present, more general Bayesian frameworks for TND studies in the presence of imperfect diagnostic tests are lacking.

Here, we propose a Bayesian approach for estimating vaccine effectiveness in TNDs that corrects for an imperfect diagnostic test and potential confounders. We show that correcting for imperfect sensitivity and specificity is important for obtaining accurate estimates and that our method captures the true vaccine effectiveness across a wide range of TND contexts. We also show how our method can be used to leverage the results of multiple parallel study sites, using the same or different diagnostic tests, to improve vaccine effectiveness inference. We provide open-source code that can be used to retrospectively analyze study data and prospectively plan studies.

## Model and Methods

### TND with known sensitivity and specificity

Study subjects in a test-negative design (TND) are patients arriving at a medical clinic with symptoms suggestive of the target disease. After recruitment, those who test positive (+) for the target disease are considered “test-positive cases” while those who test negative (−) are considered “test-negative controls.” By further stratifying the cases and controls by whether each is vaccinated (*v*) or unvaccinated (*u*), one can estimate vaccine effectiveness via the odds ratio for having the target disease between those with and without the vaccine,

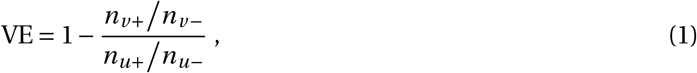

where each *n* counts individuals in the four possible vaccine and target disease status combinations.

To incorporate the fact that risk of disease can depend on patient characteristics (e.g., age and sex), an alternative approach is use a logistic regression framework in which vaccination status is one covariate among many. Letting *p*(*x*) be the probability that an individual has the target disease, letting *x*_*v*_ ∈ {0, 1} be an individual’s vaccination status, and letting *x*_1_, *x*_2_, … *x*_*m*_ be other individual covariates, the model becomes

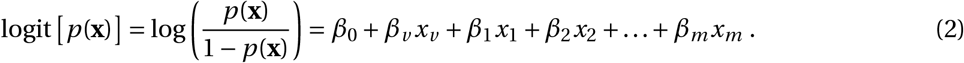

In this formulation, 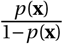 is the odds an individual with covariates **x** has the target disease and VE = 1 − exp(*β*_*v*_). The remaining *β* values reflect the impact of individual covariates on the likelihood that an individual has the target disease, thus controlling for these covariates in the estimation of vaccine effectiveness.

The above estimates assume that the diagnostic test for the target disease is reliable. However, real diagnostic tests suffer imperfect sensitivity and specificity. To account for this possibility, we model the outcome of the diagnostic test *y* for an individual with covariates **x** as the realization of a Bernoulli trial,

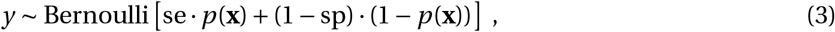

where potentially imperfect sensitivity (se) and specificity (sp) create the possibility that false negatives or false positives may affect whether *y* correctly represents the target disease state.

The TND point estimate of Eq. (1) can be adjusted for the target disease diagnostic test’s sensitivity and specificity, arriving at,

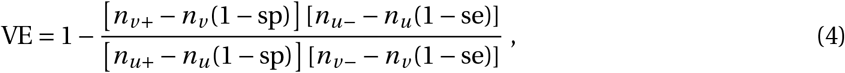

where *n*_*u*_ = *n*_*u*+_ + *n*_*u*−_ is the total number of unvaccinated individuals, and *n*_*v*_ = *n*_*v*+_ + *n*_*v*−_ is the total number of vaccinated individuals. Previous work by Endo et al. showed that Equations (1) and (4) can be subtracted to quantify the substantial estimation bias that results if one fails to correct for the diagnostic test [4].

Here we introduce a straightforward Bayesian model and logistic regression framework which simultaneously addresses the complications of (i) individual covariates, (ii) imperfect diagnostic tests, and (iii) uncertainty in sensitivity and specificity themselves. In this model, we combine the key ingredients above, by letting individual test outcomes {*y*} be given by Eq. (3), with *p*(**x**) given by Eq. (2), satisfying requirements (i) and (ii). The next section addresses how our framework satisfies requirement (iii).

### TND with uncertain sensitivity and specificity

We incorporate uncertainty about the values of sensitivity and specificity by choosing prior distributions for both parameters. In the ideal case, these priors should be anchored directly in laboratory validation data—performance on known positives and negatives—but may also be specified indirectly by analysis of point estimates from the literature. When laboratory validation data are available, in the form of *N*_neg_ known negative validation samples of which tn are correctly called true negatives, and *N*_pos_ known positive validation samples, of which tp are correctly called true positives, we model

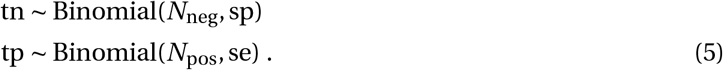

This approach, in which lab validation data directly inform both the estimates and uncertainties of the diagnostic test, follows similar joint inference models from the seroprevalence literature [9, 14]. In this study, we assume that, before seeing the test validation data, little is known about sensitivity and specificity. This assumption corresponds to using Uniform(0, 1) priors on sensitivity and specificity.

This Bayesian approach would be incomplete without specification of prior distributions on the remaining parameters. In the absence of information on the relationship between covariates and odds of infection with the target disease, we use uninformative priors for *β*_1_, *β*_2_, …, *β*_*m*_. Most importantly, for the prior distribution over the vaccine effectiveness parameter *β*_*v*_, we suggest either an uninformative prior or a prior informed by past vaccine effectiveness data, as in [6]. In this study, we used a Gamma(2, 0.5) prior for *β*_*v*_. This prior corresponds to assigning 95% probability to vaccine effectiveness being between 0.47 and 0.99 (before seeing the data). For *β*_0_, we used a Normal(0, 1) prior. This prior assigns 95% probability to the probability being between 0.12 and 0.88 that an unvaccinated individual with all other covariates equal to zero has the target disease (before seeing the data).

### Incorporating multiple diagnostic tests

A single TND study may rely on multiple diagnostic tests with potentially different sensitivities and specificities. For instance, both rapid diagnostic and PCR tests have been used to study influenza and SARS-CoV-2, with test characteristics depending on both the manufacturer and the protocol. Our Bayesian framework is easily extended to the analysis of such study designs and, similarly, permits meta-analyses of multiple studies even if each study used different diagnostic tests.

Suppose se_*j*_ and sp_*j*_ are the sensitivity and specificity of diagnostic test *j*, respectively. The test outcome *y* _*j*_ of an individual with covariates **x** who receives test *j* is modeled by

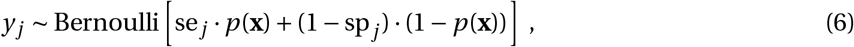

where *p*(**x**) is defined by logit [*p*(**x**)] = *β*_0_ + *β*_*v*_ *x*_*v*_ + *β*_1_*x*_1_ + … + *β*_*m*_ *x*_*m*_ as in Eq. (2).

Here, by not indexing the *β* coefficients by the test *j*, we are assuming that the various tests are being used to study either a single population or multiple populations with similar risk of disease. However, if this is not the case, this model may be extended to a hierarchical model in which *β* coefficients (except for *β*_*v*_ are indexed by the population from which the data were sampled.

Lastly, if lab validation data for sensitivity and specificity are available, such information may be incorporated separately into the model for each type of diagnostic test, via

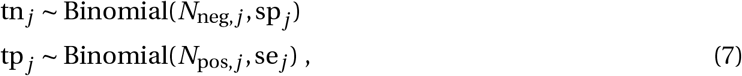

where, for test *j, N*_neg,*j*_ is the number of negative validation samples of which tn_*j*_ are correctly called true positives and *N*_pos,*j*_ is the number of positive validation samples of with tp*j* are correctly called true positives.

## Results

This section is split into two parts. The first part addresses estimating vaccine effectiveness when sensitivity and specificity are imperfect but their values are exactly known. In the Bayesian framework, this amounts to putting point priors on sensitivity and specificity. The second part addresses estimating vaccine effectiveness when sensitivity and specificity are, themselves, estimated with uncertainty from laboratory validation data. In the demonstrations that follow, we consider only one covariate but the Bayesian logistic regression approach can incorporate additional covariates as needed.

### Imperfect but known sensitivity and specificity

First, in comparison to existing vaccine effectiveness estimators, we investigated whether our corrected Bayesian vaccine effectiveness inferences are accurate across a range of TND scenarios. To do so, we simulated study data with known parameter values and then tested our ability to recover the true vaccine effectiveness from the generated data with a 95% credible interval (highest density posterior interval, HDPI) quantifying uncertainty in the vaccine effectiveness estimate. Figure 1 shows an example of one such simulation in which the corrected estimates accurately capture the true vaccine effectiveness. In this example, uncorrected estimates, which assume perfect sensitivity and specificity, show substantial downward bias, and the uncorrected 95% credible interval fails to capture the true vaccine effectiveness.

**Figure 1:**
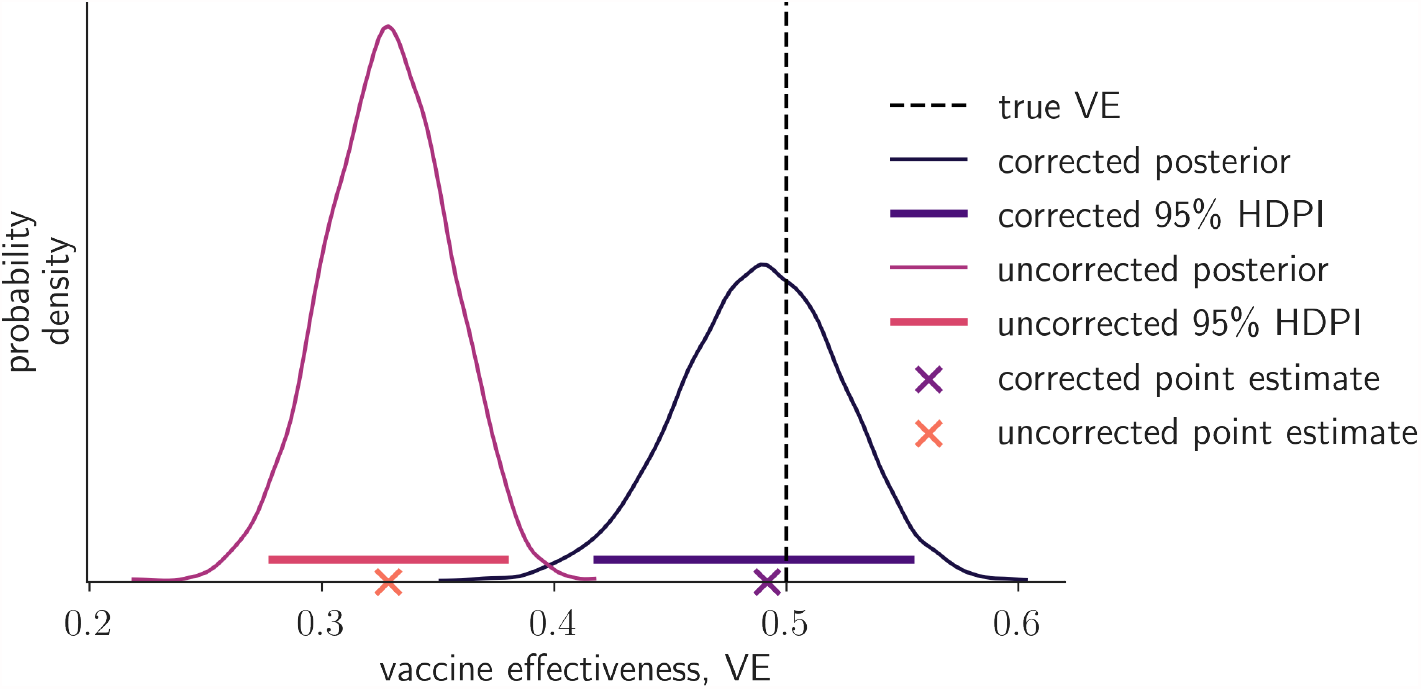
Corrected and uncorrected vaccine effectiveness point estimates, Bayesian posteriors, and 95% HDPIs. For simulated study data with true sensitivity = specificity = 0.8, the uncorrected estimates assume (incorrectly) that sensitivity and specificity are equal to one. The corrected inferences assume (correctly) that the sensitivity and specificity are 0.8. This figure illustrates the importance of adjusting for imperfect sensitivity and specificity. The simulated data used to generate this figure consisted of a vaccinated and unvaccinated group. The vaccinated group had consisted of 2060 test positive individuals and 2962 test negative individuals. The unvaccinated group consisted of 2532 test positive individuals and 2456 test negative individuals.

To more systematically evaluate the impact of test misclassification error and confirm our approach’s ability to estimate vaccine effectiveness, we created 500 synthetic datasets with known true parameter values. For each dataset, we inferred vaccine effectiveness using both the corrected and uncorrected models. To generate the 500 sets of parameter values, we drew parameters independently from the following distributions:

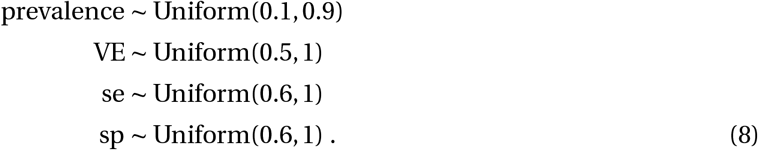

For all simulations, we assumed a study size of 5000 individual, each of whom was vaccinated with probability 0.5. Given each individual’s vaccination status *x*_*v*_, the individual’s test status was determined by a Bernoulli trial with probability

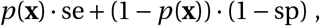

where logit[*p*(**x**)] = *β*_0_ +*β*_*v*_ *x*_*v*_ is the probability an individual with covariates **x** has the target disease from Eq. (2).

Across the 500 simulations, 97.6% (488/500) of the corrected 95% HDPIs captured the true vaccine effectiveness. The mean absolute error of the corrected point estimates was 0.036 (Fig. 2A,B). On the other hand, only 4.8% (24/500) of the uncorrected 95% HDPIs captured the true vaccine effectiveness and the mean absolute error of the uncorrected point estimates was 0.257 (Fig. 2C,D). The uncorrected estimates systematically underestimated vaccine effectiveness aligning with analytical predictions [4].

**Figure 2:**
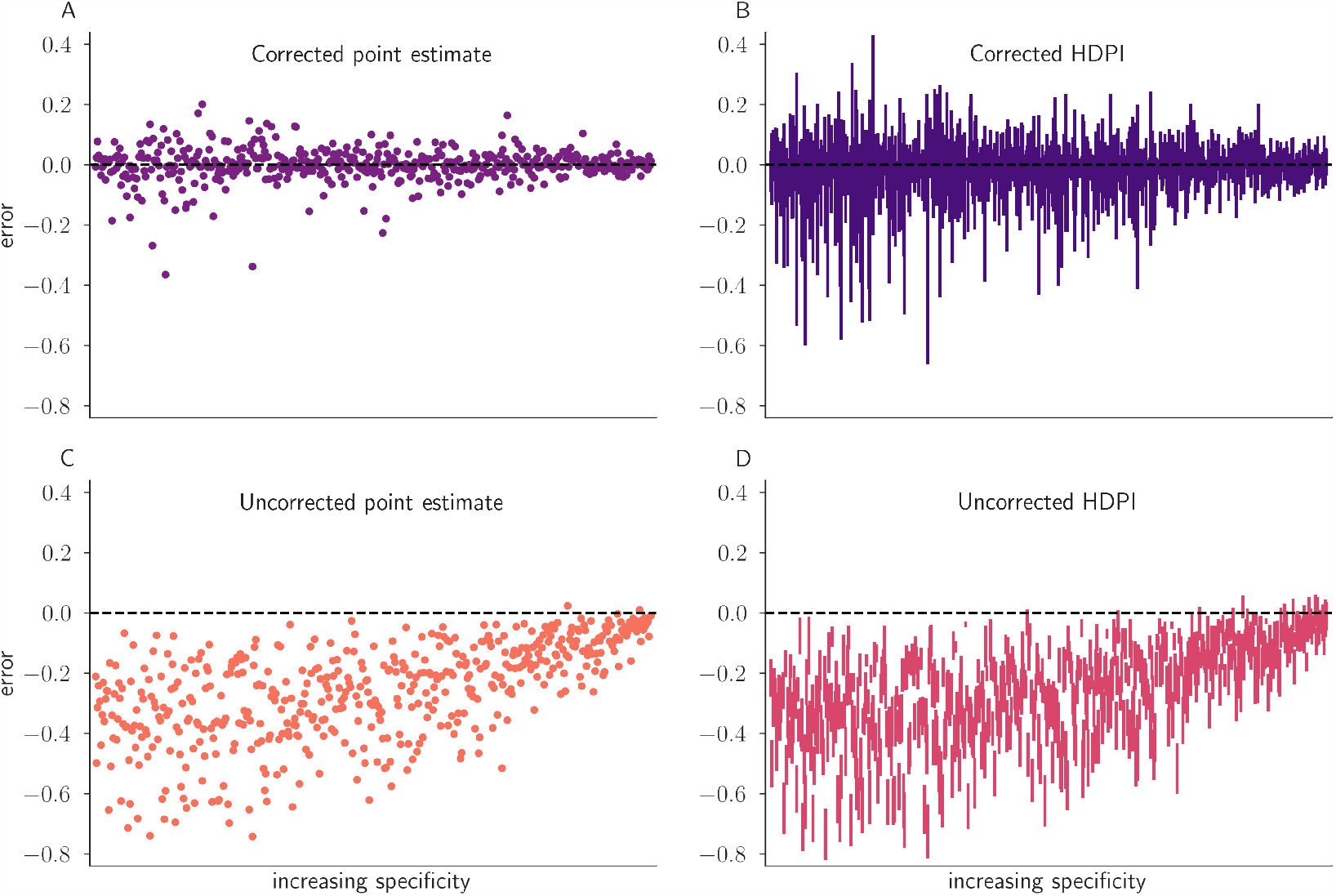
Corrected and uncorrected vaccine effectiveness point estimates and Bayesian highest density posterior intervals (HDPIs) across a variety of true parameter scenarios. Each of the 500 scenarios is characterized by four parameters: VE, prevalence, se, and sp, all drawn according to Eq. (8). Scenarios are analyzed through (A) corrected Bayesian point estimates, (B) corrected Bayesian 95% HDPIs, (C) uncorrected Bayesian point estimates, and (D) uncorrected Bayesian 95% HDPIs. Scenarios are plotted from high specificity (top) to low specificity (bottom), illustrating the relationship between uncertainty, bias, and specificity.

Imperfect tests have two further effects on inference. First, as test performance worsens, the bias of un-corrected estimates increases, revealed by plotting uncorrected estimates in order by specificity (Fig. 2C,D). Second, as test performance worsens, uncertainty of all estimates increases, as quantified by HDPI widths (Fig. 2B,D).

### Combining data from different diagnostic tests

To illustrate how evidence from multiple tests may be integrated into a single joint vaccine effectiveness estimate, we used the same simulation framework used to create Fig. 1 to simulate data from two different diagnostic tests, (i) se = 0.7 and sp = 0.95, and (ii) se = 0.5 and sp = 0.9, representing hypothetical laboratory and a rapid diagnostic tests, respectively. To generate data, *β*_0_ was set to logit(0.5) and *β*_*v*_ was set to log(0.5).

After generating data for the two datasets and performing inference on the two datasets separately and then jointly, we found that the vaccine effectiveness inference from our joint Bayesian model is more accurate (as measured by the distance between the true vaccine effectiveness and the posterior mean or median) and has less uncertainty (as measured by HDPI widths) than from either data source individually (Fig. 3). While this analysis shows how data from two different diagnostic tests can be combined to produce improved estimates over either data source on its own, the model could be extended to include additional tests as appropriate.

**Figure 3:**
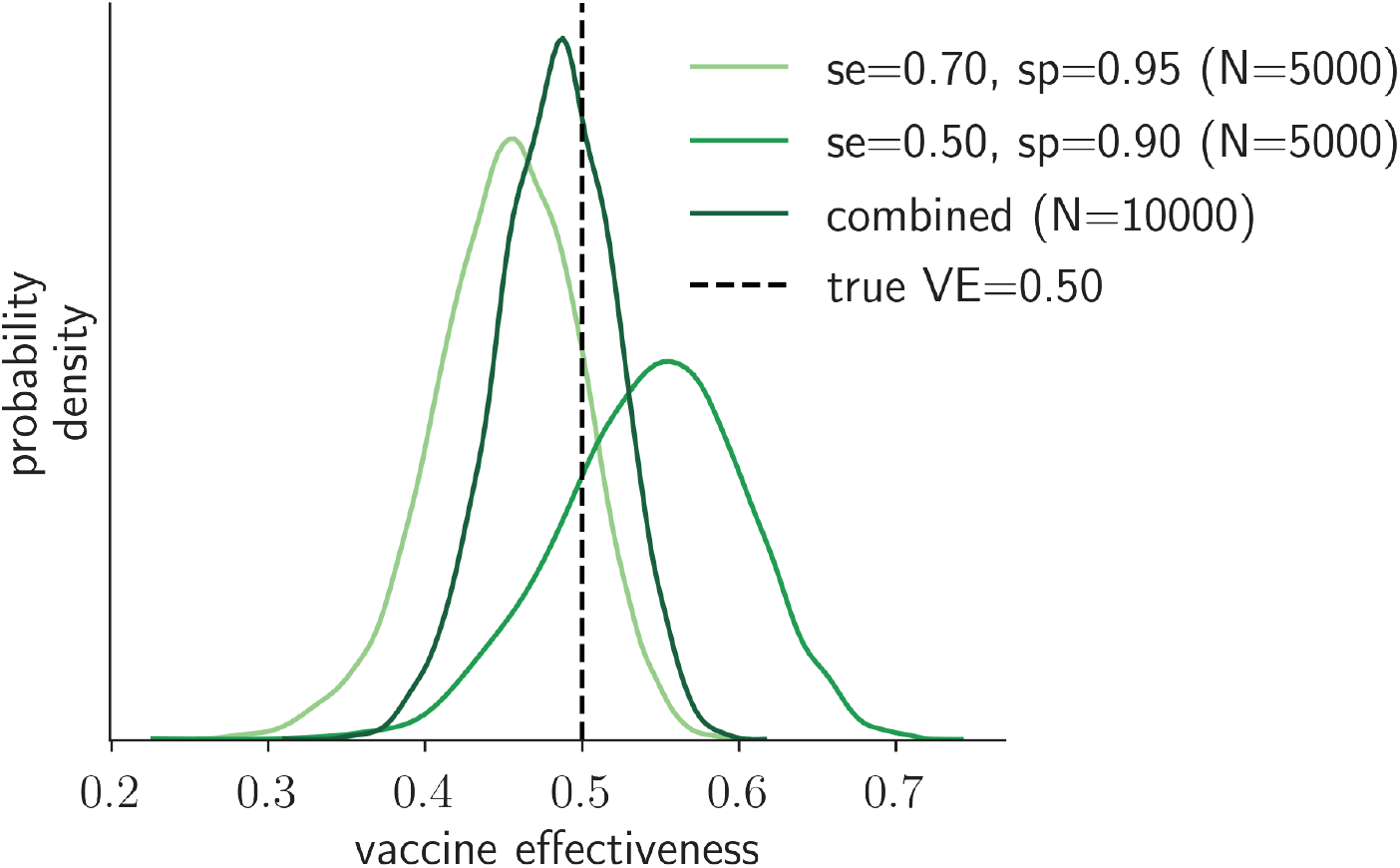
Vaccine effectiveness inferences can be improved by combining data from different diagnostic tests. For two (simulated) studies of 5000 individuals, one of which used a diagnostic test with se = 0.7 and sp = 0.95 and the other used a diagnostic test with se = 0.5 and sp = 0.90, the vaccine effectiveness posteriors are shown. Inferring vaccine effectiveness in a Bayesian framework that incorporates both studies is better than inferring vaccine effectiveness from either of the two studies separately.

### Imperfect and uncertain sensitivity and specificity

Sensitivity and specificity are typically estimated from limited validation data [9, 13, 14] and, thus, they carry uncertainty. Failure to account for this uncertainty may lead to erroneous inferences and conclusions [8, 9]. In this section, we examine how our Bayesian framework propagates uncertainty in sensitivity and specificity into vaccine effectiveness inferences.

First, we examined how increasing laboratory validation efforts leads to decreasing uncertainty in sensitivity and specificity, in turn affecting vaccine effectiveness inferences. If sensitivity is estimated via tp true positive results in *N*_pos_ tests of known positive individuals and specificity is estimated via tn true negative results in *N*_neg_ tests of known negative individuals, then the model is

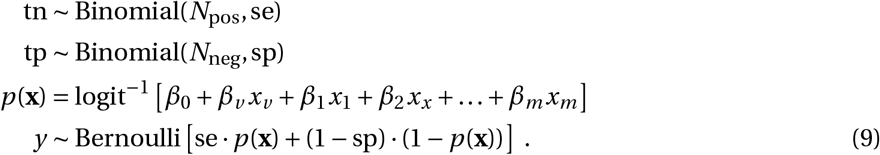

We assigned uniform priors to sensitivity and specificity, equivalent to assuming that all values are equally plausible before seeing the validation data.

We explored how using a validation study to estimate sensitivity and specificity affects vaccine effectiveness inference by using the model in Eq. (9) to simulate data. For simulations, we set *N* = 5000, se = 0.7, sp = 0.95, *β*_0_ = logit(0.5), and *β*_*v*_ = log(0.5) and we varied the amount of data used to estimate sensitivity and specificity, *N*_neg_ and *N*_pos_, ranging from 0 validation samples to 1000 valudation samples. After simulating data, we then used the Bayesian model in Eq. (9) to infer the parameters that generated the data. Since the model in Eq. (9) jointly models sensitivity, specificity, and vaccine effectiveness (through *β*_*v*_), the Bayesian posterior allows us to estimate sensitivity and specificity in addition to vaccine effectiveness.

As the amount of data used to estimate sensitivity and specificity increases (i.e., *N*_pos_ and *N*_neg_ increase), uncertainty in their values decreases accordingly (Fig. 4A,B), thereby improving vaccine effectiveness inference (Fig. 4C). As expected, increasing the amount of data used to estimate sensitivity and specificity causes the posteriors for sensitivity and specificity to tighten substantially around their true values, while the posterior for vaccine effectiveness tightens modestly, reflecting combined uncertainties from both validation and field data.

**Figure 4:**
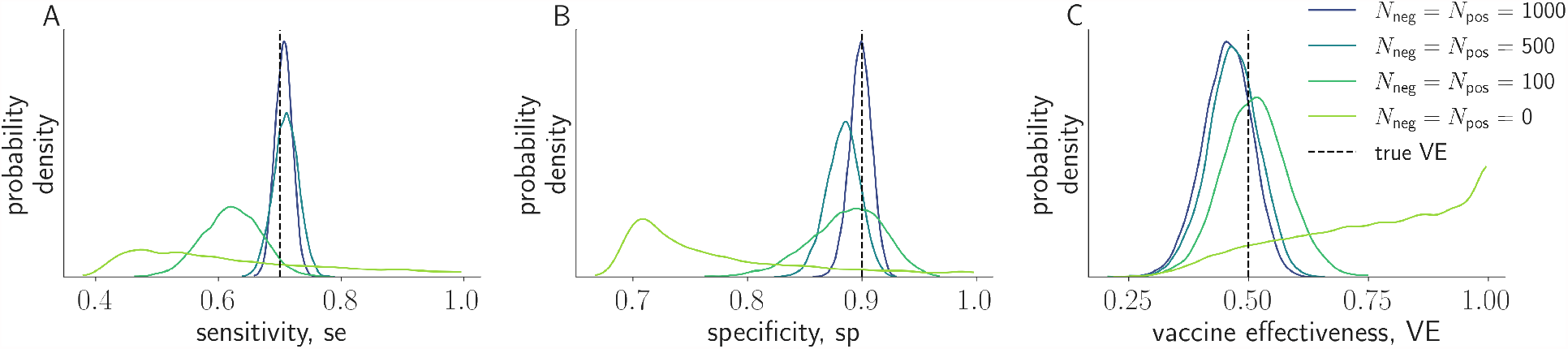
Increasing the amount of data used to validate sensitivity and specificity improves inferences. The posteriors for different number of tests used to validate sensitivity and specificity are shown. Here, num tests = *N*_pos_ = *N*_neg_. We show the posteriors for se (A), sp (B), and VE (C). As the number of tests used to validate sensitivity and specificity increases, the posteriors tighten around the true parameter values.

Previous work has shown that vaccine effectiveness estimates are more affected by poor sensitivity in some scenarios and poor specificity in others [3, 4], thereby informing test and test cutoff selection. We investigated a related question by asking whether vaccine effectiveness estimates are more affected by low precision in sensitivity vs specificity estimates. To investigate the potential trade-off between precision in sensitivity versus specificity, we fixed the total number of diagnostics tests used to validate sensitivity and specificity (i.e., we fixed *N*_neg_ + *N*_pos_), and varied the proportion of the total that went toward validating specificity as opposed to sensitivity.

We simulated test validation and vaccine effectiveness TND data via Eq. (9) with fixed *N*_neg_ + *N*_pos_ = 400. We assumed Uniform(0, 1) priors on se and sp. For each set of true parameters (i.e., the relative prevalence of the target disease, the probability that an individual is vaccinated, vaccine effectiveness, sensitivity, and specificity), we varied the proportion of the total number of validation tests that went to estimating specificity. And, for each proportion, we performed 50 simulations and averaged the vaccine effectiveness 95% HDPIs (Fig. 5A) and the bias of the vaccine effectiveness posterior mean (Fig. 5B).

**Figure 5:**
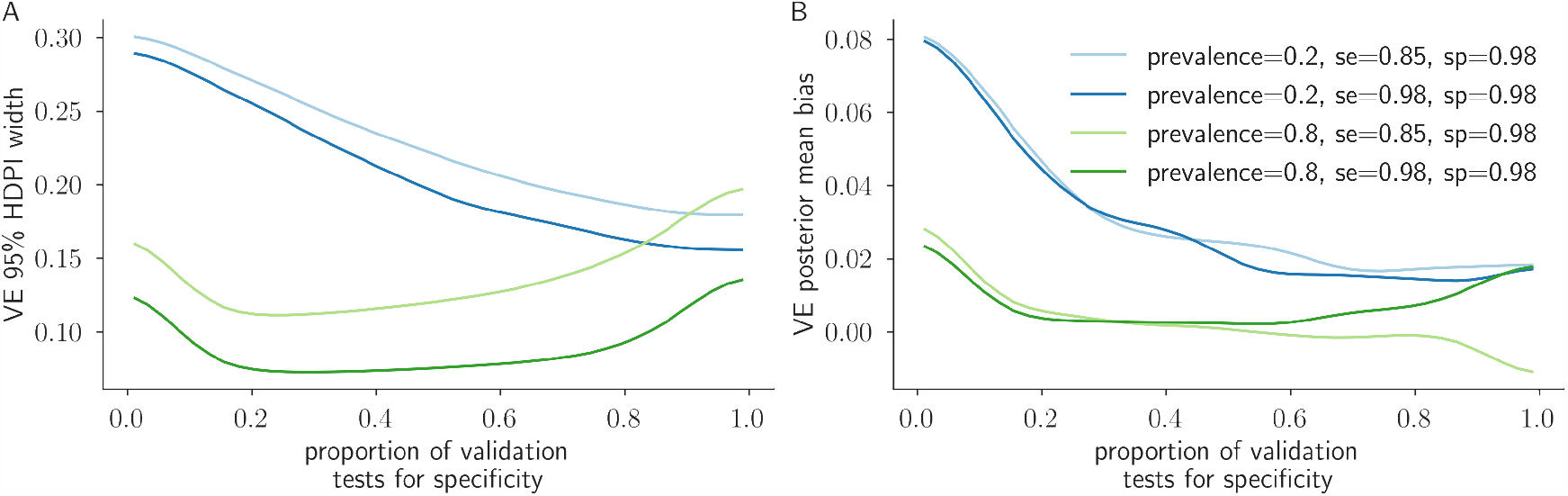
Determining the best allocation of a fixed number of validation tests. The horizontal axis corresponds to the proportion of 400 diagnostic tests that were allocated to validating specificity. For each proportion, sensitivity and specificity validation studies were simulated along with a vaccine effectiveness (VE) study of 5000 individuals. Independent uniform priors on the interval [0, 1] were used on test sensitivity and specificity before seeing the validation data. We fixed vaccine effectiveness and the probability a study participant is vaccinated at 0.75 and 0.5, respectively. The true relative prevalence of the target disease (prevalence), sensitivity (se), and specificity (sp) were varied to generate the curves shown. For each curve and each proportion of validation tests used to validate specificity, we conducted 50 simulations. Over these 50 simulations, we computed the average vaccine effectiveness 95% HDPI (A) and posterior mean bias (B). The curves show how one can allocate validation tests to maximally benefit vaccine effectiveness inference (i.e., to produce 95% HDPIs of minimal width and bias).

To estimate vaccine effectiveness with minimal uncertainty and bias, we found that the relative importance of precision in sensitivity versus specificity depends on the scenario (e.g., the target disease’s relative prevalence) (Fig. 5). In low relative prevalence scenarios in which there are few true positives, it is generally more beneficial to precisely estimate specificity. In high relative prevalence scenarios in which there are few true negatives, it is generally more beneficial to precisely estimate sensitivity (Fig. 5). More broadly, this experiment illustrates how one may investigate the effect of both the *values* of sensitivity and specificity and the *precision* in their values on vaccine effectiveness inference.

Finally, validation data contain information about sensitivity and specificity and, thus, having validation data updates the uninformative Uniform(0, 1) priors on sensitivity and specificity. However, the TND data also reflects information about sensitivity and specificity. As a consequence, even if all the validation tests are allotted to validating sensitivity, the posterior on specificity will not be Uniform(0, 1) but will reflect the limited inferences about specificity that can be drawn from TND data (Fig. S2).

## Discussion

This study introduced and demonstrated a Bayesian approach to vaccine effectiveness estimation via a test-negative design, adjusted for imperfect test sensitivity and specificity as well as individual covariates. Our approach avoids the pitfalls of bias, by adjusting for sensitivity and specificity, and of false precision, by incorporating the uncertainty associated with sensitivity and specificity themselves. Based on this framework, we showed how data from both the field site and validation labs may be used to produce joint posterior estimates of vaccine effectiveness, sensitivity, and specificity. This allowed us to demonstrate how vaccine effectiveness estimates may be made more precise simply by better validating the performance characteristics of diagnostic tests. We further showed how our Bayesian model can be used to estimate vaccine effectiveness from multiple data streams at once, each based on a different diagnostic test.

One important finding is that imperfect sensitivity and specificity lead to increased uncertainty of vaccine effectiveness estimates, even when vaccine effectiveness estimates are corrected and unbiased (Fig. 2). Thus, while several preceding studies have shown that estimates can be made unbiased by adjusting for sensitivity and specificity [5, 3, 4], here we further showed that test performance affects vaccine effectiveness uncertainty even after bias adjustment, leading us to conclude that there is independent value in high-performance diagnostic testing, even if signals may be statistically debiased *a posteriori*.

Uncertainty is also affected by sample size, both for the number of individuals recruited into the study as well as the number of negative and positive controls used to validate test performance. Our approach and accompanying software allow one to explore various choices for these sample sizes, linking them to uncertainty of vaccine effectiveness inferences measured via the width of the vaccine effectiveness posterior credible interval. This, in turn, allows for decisions as to whether or not a larger study or additional test validation efforts are likely to be worth the additional effort (Fig. 4).

As with any statistical model, frequentist or Bayesian, our model makes assumptions which should be highlighted. One assumption is that there is no misclassification of vaccination status (i.e. exposure misclassification). This assumption has been relaxed in some applications others [5, 10, 15, 16, 17, 18]. We also assumed that sensitivity and specificity are independent, yet some assays featuring cutoff parameters necessarily incur a sensitivity-specificity tradeoff, which may require additional care [15, 16].

With increasing emphasis being placed on reproducibility and open data practices, our work highlights the importance of reporting the raw outcomes of validation tests, not just point estimates. These values have meaningful impacts on not only the uncertainty of estimates [14], but study conclusions as well [9].

## Data Availability

All code needed to evaluate the conclusions in the paper are present in the paper and/or the Supplementary Materials, and open-source code is available at https://github.com/erikj540/Bayesian-Vaccine-Efficacy. The Bayesian models were implemented in the programming language STAN which we called through Python 3.8.

## Acknowledgements

This work was supported in part by the SeroNet program of the National Cancer Institute (1U01CA261277-01), and by the Morris-Singer Fund for the Center for Communicable Disease Dynamics at the Harvard T.H. Chan School of Public Health.

## Code Availability

All code needed to evaluate the conclusions in the paper are present in the paper and/or the Supplementary Materials, and open-source code is available^1^. The Bayesian models were implemented in the programming language STAN which we called through Python 3.8.

## Ethics Declaration

R.K. received consulting fees from Partners In Health. M.L. received grants from US CDC, NIH/NIGMS, NIH/NIAID, and UK National Institute for Health Research, and Pfizer, and consulting fees or honoraria from Merck, Sanofi Pasteur, Janssen, and Bristol Myers Squibb. E.K.J., Y.H.G., and D.B.L. declare no competing interests.

## Supplementary Material

**Figure S1:**
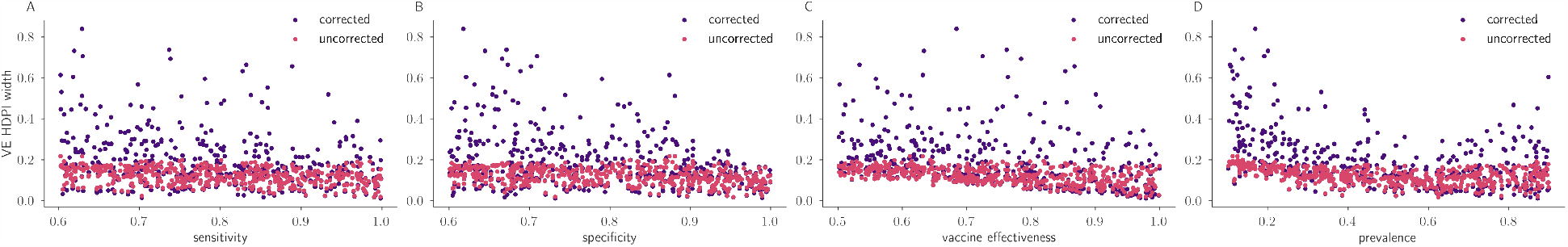
Vaccine effectiveness (VE) highest posterior density interval (HDPI) widths as a function of increasing parameter values. For the same simulated scenarios used to generate Fig. 2, we plot the corrected and uncorrected HDPI widths (corrected in purple, uncorrected in pink) as functions of the four simulation parameters: (A) sensitivity, (B) specificity (as in Fig. 2), (C) vaccine effectiveness, and (D) prevalence which is, more specifically, the relative prevalence of the target disease. The uncorrected HDPIs generally have narrower HDPIs as they assume sensitivity and specificity are perfect (i.e., they assume se = sp = 1) which leads to erroneously confident (i.e., narrow) credible intervals. As specificity nears one, the HDPI widths become narrow reflecting the importance of high specificity in estimating vaccine effectiveness. Interestingly, as a function of prevalence, the vaccine effectiveness HDPI is narrowest when the target disease is approximately equally as prevalent as the non-target diseases, i.e., when prevalence ≈ 0.5 (Fig. S1D).

**Figure S2:**
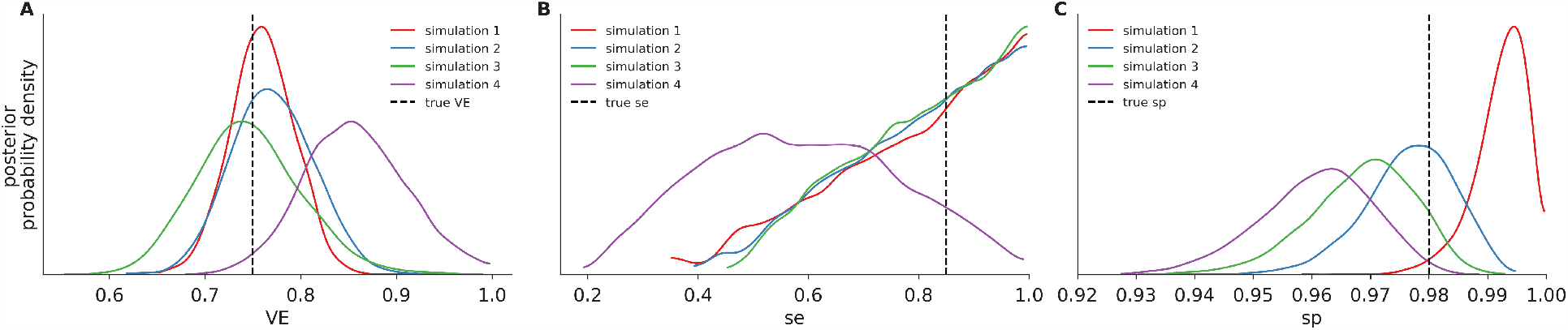
Vaccine effectiveness (VE), sensitivity (se), and specificity (sp) posteriors. Here we show vaccine effectiveness (A), sensitivity (se), and specificity (sp) posteriors for four randomly selected simulations of the 50 simulations used to produce Fig. 5 with true parameter values: relative prevalence= 0.2, se = 0.85, sp = 0.98, and the proportion of the 400 validation tests used to validate specificity= 0.99. Although the vast majority of the validation tests were used to validate specificity, the vaccine effectiveness TND data contains information about sensitivity that our Bayesian inference framework uses to update the Uniform(0, 1) prior.

## Footnotes

1 https://github.com/erikj540/Bayesian-Vaccine-Efficacy

## References

[1] Wakaba Fukushima and Yoshio Hirota. Basic principles of test-negative design in evaluating influenza vaccine effectiveness. Vaccine, 35(36):4796–4800, 2017.

[2] Huiying Chua, Shuo Feng, Joseph A Lewnard, Sheena G Sullivan, Christopher C Blyth, Marc Lipsitch, and Benjamin J Cowling. The use of test-negative controls to monitor vaccine effectiveness: a systematic review of methodology. Epidemiology (Cambridge, Mass.), 31(1):43, 2020.

[3] Michael L Jackson and Kenneth J Rothman. Effects of imperfect test sensitivity and specificity on observational studies of influenza vaccine effectiveness. Vaccine, 33(11):1313–1316, 2015.

[4] Akira Endo, Sebastian Funk, and Adam J Kucharski. Bias correction methods for test-negative designs in the presence of misclassification. Epidemiology & Infection, 148, 2020.

[5] Tom De Smedt, Elizabeth Merrall, Denis Macina, Silvia Perez-Vilar, Nick Andrews, and Kaatje Bollaerts. Bias due to differential and non-differential disease-and exposure misclassification in studies of vaccine effectiveness. PLoS One, 13(6):e0199180, 2018.

[6] Michael L Jackson, Jill Ferdinands, Mary Patricia Nowalk, Richard K Zimmerman, Burney Kieke, Manjusha Gaglani, Kempapura Murthy, Joshua G Petrie, Emily T Martin, Jessie R Chung, et al. Differences between frequentist and bayesian inference in routine surveillance for influenza vaccine effectiveness: a test-negative case-control study. BMC public health, 21(1):1–8, 2021.

[7] Joseph A Lewnard, Manish M Patel, Nicholas P Jewell, Jennifer R Verani, Miwako Kobayashi, Mark W Tenforde, Natalie E Dean, Benjamin J Cowling, and Benjamin A Lopman. Theoretical framework for retrospective studies of the effectiveness of SARS-CoV-2 vaccines. Epidemiology, 32(4):508–517, 2021.

[8] Eran Bendavid, Bianca Mulaney, Neeraj Sood, Soleil Shah, Emilia Ling, Rebecca Bromley-Dulfano, Cara Lai, Zoe Weissberg, Rodrigo Saavedra-Walker, Jim Tedrow, et al. COVID-19 antibody seroprevalence in Santa Clara County, California. MedRxiv, 2020.

[9] Andrew Gelman and Bob Carpenter. Bayesian analysis of tests with unknown specificity and sensitivity. Journal of the Royal Statistical Society: Series C (Applied Statistics), 69(5):1269–1283, 2020.

[10] Paul Gustafson, Nhu D Le, and Refik Saskin. Case–control analysis with partial knowledge of exposure misclassification probabilities. Biometrics, 57(2):598–609, 2001.

[11] James R Marshall. The use of dual or multiple reports in epidemiologic studies. Statistics in medicine, 8(9):1041–1049, 1989.

[12] Neal D Goldstein, Igor Burstyn, E Claire Newbern, Loni P Tabb, Jennifer Gutowski, and Seth L Welles. Bayesian correction of misclassification of pertussis in vaccine effectiveness studies: How much does underreporting matter? American journal of epidemiology, 183(11):1063–1070, 2016.

[13] Daniel B Larremore, Bailey K Fosdick, Kate M Bubar, Sam Zhang, Stephen M Kissler, C Jessica E Metcalf, Caroline O Buckee, and Yonatan H Grad. Estimating sars-cov-2 seroprevalence and epidemiological parameters with uncertainty from serological surveys. Elife, 10:e64206, 2021.

[14] Daniel B Larremore, Bailey K Fosdick, Sam Zhang, and Yonatan H Grad. Jointly modeling prevalence, sensitivity and specificity for optimal sample allocation. bioRxiv, 2020.

[15] Matthew P Fox, Timothy L Lash, and Sander Greenland. A method to automate probabilistic sensitivity analyses of misclassified binary variables. International journal of epidemiology, 34(6):1370–1376, 2005.

[16] Haitao Chu, Zhaojie Wang, Stephen R Cole, and Sander Greenland. Sensitivity analysis of misclassification: a graphical and a bayesian approach. Annals of Epidemiology, 16(11):834–841, 2006.

[17] Richard F MacLehose, Andrew F Olshan, Amy H Herring, Margaret A Honein, Gary M Shaw, Paul A Romitti, et al. Bayesian methods for correcting misclassification an example from birth defects epidemiology. Epidemiology (Cambridge, Mass.), 20(1):27, 2009.

[18] George Luta, Melissa B Ford, Melissa Bondy, Peter G Shields, and James D Stamey. Bayesian sensitivity analysis methods to evaluate bias due to misclassification and missing data using informative priors and external validation data. Cancer epidemiology, 37(2):121–126, 2013.

